# Bibliometric analysis of parental anxiety and postpartum depression across the perinatal period from 1920-2020: A protocol

**DOI:** 10.1101/2021.05.08.21256829

**Authors:** Justine Dol, Marsha Campbell-Yeo, Cindy-Lee Dennis, Patricia Leahy-Warren

## Abstract

**Introduction:** Throughout the perinatal period from pregnancy to the first year postpartum, both men and women experience significant physical, psychological, and social changes which may increase their risk of a mental illness, including anxiety and depression. There has been significant growth in the frequency literature around anxiety and depression across the perinatal period over the past decades with significant variation in definition, measurement outcomes, and populations. To focus future research and identify gaps, it is important to explore current patterns and trends in the current literature.

**Objective:** The objective of this bibliometric analysis is to analyze the characteristics and trends in published research on anxiety and depression across the perinatal period from January 1, 1920 to end of 2020.

**Inclusion criteria:** All published literature in Web of Science on perinatal anxiety and depression from January 1, 1920 to December 31, 2020.

**Methods:** Web of Science will be used to analyze bibliometric information through their built-in analysis feature and citation report that generates a list of leading publications, publication years, document types, authors, source titles, countries/regions, organizations, and research areas. VOSViewer will be utilized to analyze and visualize the networks of linkages between the identified reports, including bibliometric networks, including co-authorship, co-occurrence, and co-citation, as well as co-occurrence between keywords.

**Conclusion:** The findings from this study will provide useful information to guide future work on perinatal anxiety and depression. This bibliometric review will provide an overview of the work to date in perinatal mental health, identify key contributions to the field, and identify knowledge gaps and future directions.

## INTRODUCTION

Throughout the perinatal period from pregnancy to the first year postpartum, mothers and fathers experience significant physical, psychological and social changes which may increase their risk of a mental illness, including anxiety and depression (O’Hara & Wisner, 2014). Over the years, there has been a growth in literature on perinatal mental illness and in 1994, postpartum depression was recognized in the Diagnostic and Statistical Manual (DSM-IV) as a Major Depressive Disorder with Postpartum Onset (American Psychiatric Association, 2013). While there is no specific recognition in the DSM-V of anxiety specific to the perinatal experience, anxiety in all populations is the most common mental illnesses diagnosed each year (Ströhle et al., 2018).

Perinatal depression is a non-psychotic depressive episode that begins in or extends from pregnancy into the postpartum period, ranging from minor depressive symptoms to a clinical diagnosis (Lanes et al., 2011). Symptoms can include anxiety, guilt, negative maternal attitudes and attachment, poor parenting self-efficacy and coping skills, lasting up to 14 months postpartum (Lanes et al., 2011). While often comorbid with perinatal depression, perinatal anxiety is a separate mental health concern, shaped by fear and worry, rather than depressive thoughts, and can emerge as generalized anxiety, panic disorders, obsessive compulsive disorder, or post-traumatic stress disorder (Ali, 2018). Anxiety can manifest itself as several symptoms, including “excessive and persistent fear, worry, and tension and regularly includes physical symptoms such as sleeping difficulties and inability to concentrate. Severe symptoms include panic and recurrent intrusive thoughts or images, often related to the harm of their child” (Dennis et al., 2016, p. 486). While anxiety and depression may be pre-existing or may emerge in the antenatal period, studies have shown that it is important to re-assess for possible anxiety and depression in the postpartum period due to the changes that occur after birth and variation in symptoms across the perinatal period (Andersson et al., 2006; Bayrampour et al., 2016).

There has been significant growth in the literature around depression and anxiety across the perinatal period over the past decades, with significant variation in definition and subsequent measurements of these mental health outcomes (Meades & Ayers, 2011; Sinesi et al., 2019; Ukatu et al., 2018) as well as co-reported psychosocial outcomes (Leach et al., 2017; Norhayati et al., 2015; Wee et al., 2011). To focus future research and identify gaps, it is important to explore current patterns and trends in the current literature. To do this, we will use a bibliometric analysis approach which seeks to comprehensively explore patterns in publications in a given research area, including trends over time and the influences of contributions by citations, such as relationships between authors and publications (Donthu et al., 2021; Gauthier, 1998). Bibliometric analyses has been used across a variety of academic disciplines to provide a broad overview of the current literature to provide description, evaluation, and scientific and technological monitoring (Donthu et al., 2021; Gauthier, 1998). While there have been many reviews on anxiety and depression across the perinatal period, none to date have used this approach to map the evolution of published literature on perinatal mental health to characterize research outputs, distribution, and relationships.

The objective of this bibliometric analysis is to analyze the characteristics and trends in published research on anxiety and depression across the perinatal period over the past hundred years, from January 1, 1920 to December 31, 2020. Specifically, this study aims to identify, analyze, and visualize research publications on perinatal anxiety and depression, exploring outcomes and co-relations such as author, country, institution, and year to provide an overview on perinatal mental health research over the last hundred years.

## METHODS

This study follows the standard bibliometric approach (Donthu et al., 2021; Gauthier, 1998; Linnenluecke et al., 2020), using the following steps: “(1) define the search criteria, keywords, and time periods; (2) selection of Web of Science database; (3) adjustment and refinement of research criteria; (4) full export of result; (5) analysis of the information and discussion of the results” (Ruiz-Real et al., 2018, p. 2).

A preliminary search of MEDLINE, the Cochrane Database of Systematic Reviews and PROSPERO was conducted and no current or underway systematic reviews, scoping reviews, or bibliometric analysis on the topic were identified.

### Citation Database

The search strategy will be conducted in Web of Science using the Web of Science Core Collection. This includes Science Citation Index Expanded (SCI-EXPANDED) --1900-present, Social Sciences Citation Index (SSCI) --1956-present, Arts & Humanities Citation Index (A&HCI) --1975-present, Conference Proceedings Citation Index- Science (CPCI-S)--1990-present, Conference Proceedings Citation Index- Social Science & Humanities (CPCI-SSH)--1990-present, and Emerging Sources Citation Index (ESCI) --2015-present.

The use of a singular database is recommended in bibliometric reviews and may vary depending on question of interest (Donthu et al., 2021; Ruiz-Real et al., 2018). For the purpose of this review, Web of Science was selected as the only database as it: (1) has 90 million records from 256 disciplines from 1900 to present, representing a good breadth of relevant citations; (2) provides a summary report on key bibliometric outcomes including publication, authors, and research areas; and (3) exports into a format which is required for analysis in software tool, VOSviewer (Moral-Muñoz et al., 2020). While Scopus was considered as an alternative that is commonly used in bibliometric analyses, it was not selected as it has a total limit download of any query of 2,000 report. A preliminary search of the term “postpartum depression” yielded 5,030 records in Scopus, thus deeming it not conducive to the current review. Finally, when the search was run in PubMed, Scopus, PsycInfo, and Web of Science, Web of Science had the largest number of records identified, thus solidifying it as the ideal sole database for this topic despite potential limitations (e.g., citation counts may vary from other databases, etc.).

### Search Strategy

The search strategy has been developed by leading experts in the perinatal mental health field in consultation with a health science librarian using terms for postpartum depression and anxiety as used in the lireature. Separate searches were developed for anxiety and depression and are available in Table 1.

**Table 1.**
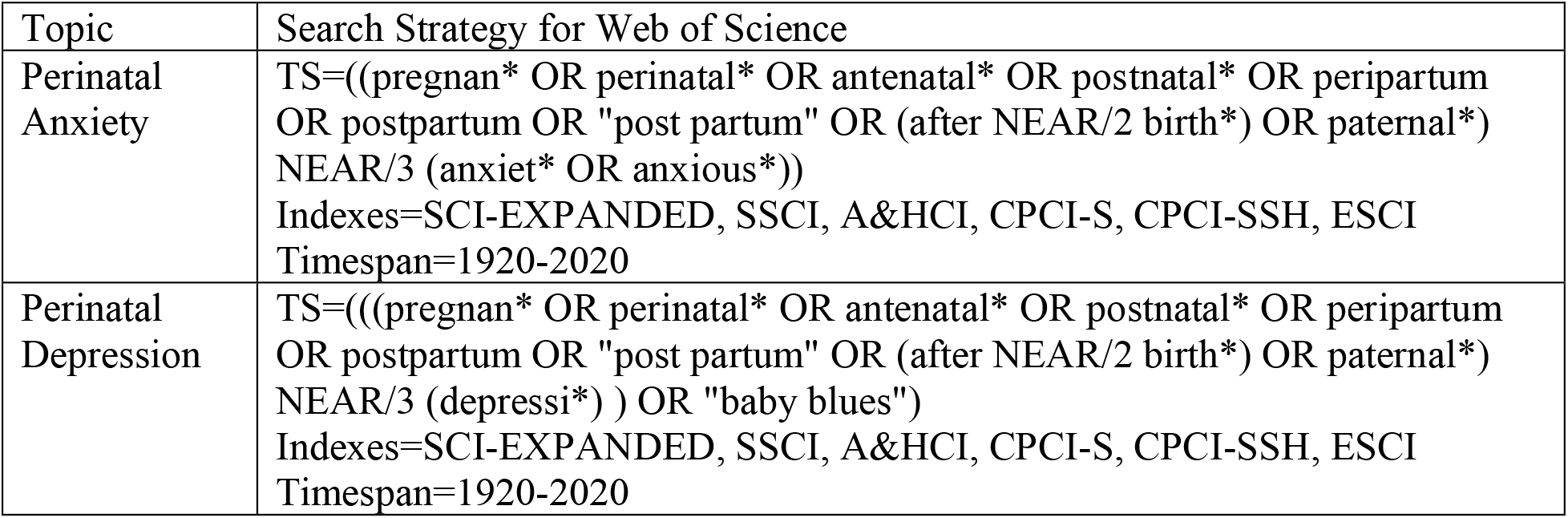
Search strategy used for the bibliometric review on perinatal anxiety and depression

### Inclusion and Exclusion Criteria

Literature will be considered for inclusion published since January 1, 1920 to December 31, 2020 published in any language. The following exclusion criteria will be applied and the number of studies that were excluded will be identified in the full report:

- Records in 2021 will be excluded as the records here would not have had time to be cited and it would create an incomplete picture in the timeline as it only reflects a partial year of publications.
- Records must be classified either an article or review, excluding conference abstracts, books/book reviews, and other non-peer reviewed material.

### Procedure

Once the search is run, the outputs will be exported with all available information, including Full Records and Cited References, in “.txt” format, which will be used for the bibliometric analysis in VOSviewer, as well as “.csv” format. Due to limitations in Web of Science, reports will be downloaded in batches of 500 and merged into one singular file for analysis. Separate reports will be created for bibliometric analysis related to perinatal anxiety and depression.

## Data Analysis

### Bibliometric Data

Web of Science will be used to analyze bibliometric information through their built-in analysis feature and citation report, including authors names and number of publications per author, countries (based on author affiliations) and number of publications per country, journals and number of publications per journal, publication type and number of publications by type, year and number of publications per year, and research area (defined by Web of Science) and number of publications per research area. Additionally, the top 20 reports with the highest number of citations will be reported. Web of Science output files will be downloaded for analysis.

### Visualization Mapping

VOSViewer is a software tool that was designed specifically for bibliometric reviews and can provide visual analysis based on bibliometric networks (Moral-Muñoz et al., 2020). It will be utilized to analyze and visualize the networks of linkages between the identified reports, including bibliometric networks, including co-authorship, co-occurrence, and co-citation, as well as co-occurrence between keywords (excluding non-relevant or generic terms) (van Eck & Waltman, 2020). While networks can be generated using a variety of bibliographic database files (e.g., Web of Science, Scopus, PubMed, RIS, EndNote, etc), data from reference managers cannot be used for some analysis, including identifying citation, bibliographic coupling, and co-citation links between items (van Eck & Waltman, 2020). Therefore, using the citation output directly from Web of Science provides the most effective option to produce visualization.

Maps are created using one type of item (i.e., reports, authors, terms) which explores links, or connections, between other items, with each link having a strength (van Eck & Waltman, 2020). For co-authorship links, for example, the strength of the link would be based on the number of publications two authors have co-authored (van Eck & Waltman, 2020). A network visualization map is comprised of items (circles) and links (lines and closeness) together (van Eck & Waltman, 2020). Within the network may exist clusters, which are grouped based on similarity and items may only belong to one cluster (van Eck & Waltman, 2020). In addition, each item may have a weight or score assigned to it, with higher weights/scores regarded as more important that lower, which are shown more notably on network maps by a larger circle (van Eck & Waltman, 2020). Also possible are overlap visualization, which is the same as network visualization but are colored differently based on scores or user-defined colors, as well as density visualization, which is also similar network visualization but the color indicates density of items at that point, which higher weights being more yellow and lower weight being more blue (van Eck & Waltman, 2020).

For the current review, visualizations will be created for co-authorship (of authors and countries), keyword co-occurrence, citation (number of times reports and authors cited together), bibliographic coupling (number of shared references for reports and authors), and co-authors (number of times references, sources, and first authors cited together). Additionally, co-occurrence of keywords will be conducted based on text data using title and abstract data. Prior to this, data cleaning will occur through the identification of similar words (e.g., same author but with first initial or all initials, behaviour/behavior) and exclusion of non-relevant terms (e.g., conclusion, method, result), which will be managed through VOSViewer’s thesaurus function.

## CONCLUSION

This bibliometric review will provide insight into the publication trends in perinatal mental health over the past century, including identifying past (e.g., through co-citation analysis), present (e.g., bibliographic coupling), and future (e.g., co-word analysis) directions (Donthu et al., 2021). This bibliometric review will provide an overview of the work to date in perinatal mental health, identify key contributions to the field, and identify knowledge gaps and future directions.

## Data Availability

This is a protocol, no data is available.

## Notes

### Competing Interest Statement

The authors have declared no competing interest.

### Funding Statement

No external funding has been received.

### Author Declarations

Not applicable - protocol for a bibliometric review.

### Summary of Updates

Significant revisions have been made to the protocol for clarity and following with standard bibliometric methodology.

